# Proton pump inhibitors use and risk of preeclampsia

**DOI:** 10.1101/2021.09.15.21263602

**Authors:** Salman Hussain, Ambrish Singh, Benny Antony, Jitka Klugarová, Miloslav Klugar

**Author notes:** Corresponding author: Dr. Miloslav Klugar*, Czech National Centre for Evidence-Based Healthcare and Knowledge Translation, (Cochrane Czech Republic, Czech EBHC: JBI Centre of Excellence, Masaryk University GRADE Centre), Institute of Biostatistics and Analyses, Faculty of Medicine, Masaryk University, Brno, Czech Republic.

## Abstract

Preeclampsia is one of the common complications of pregnancy and is characterized by high blood pressure. Proton pump inhibitors (PPIs) are commonly used for the management of gastroesophageal reflux disease among pregnant women. Recently, multiple epidemiological studies suggested the association between PPIs use and the risk of preeclampsia. This study aims to review the evidence and meta-analyse the pooled risk of preeclampsia in PPI users from epidemiological studies. Databases-MEDLINE, Embase, Scopus, Web of Science Core Collection, Emcare, and CINAHL (EBSCO) as well as sources of grey literature, ProQuest Dissertations & Theses Global, ClinicalTrials.gov and WHO International Clinical Trials Registry Platform will be searched to identify the epidemiological studies assessing the association between PPIs use and the risk of preeclampsia. Study selection, data extraction, and quality assessment will be performed by two independent authors. The risk of bias among included studies will be evaluated by using the Newcastle-Ottawa scale. The pooled effect of PPIs use on the risk of preeclampsia in pregnant women is the primary outcome of interest. Meta-analysis will be performed using Review Manager version 5.4.

## Background and rationale

Preeclampsia is one of the common complications of pregnancy and is characterized by high blood pressure.^1^ It is also one of the leading causes of maternal morbidity and mortality globally.^2^ Proton pump inhibitors (PPIs) are the most commonly and widely used drug and sold as over-the-counter (OTC) in many countries.^3^ In the last decade safety of PPIs has been a matter of scrutiny. Previously, Our previous systematic reviews and meta-analyses have found PPI use to be linked with several adverse health outcomes.^4-6^ Nevertheless, PPIs are known to have an acceptable safety profile in pregnant women, and evidence from published studies reports no increased risk of congenital defects or preterm delivery in PPIs users.^7,8^ In a recent meta-analysis, no association was observed for maternal PPI use and neonatal death or preterm birth.^9^

Evidence from preclinical studies found a protective effect of PPIs in the management of preeclampsia.^10,11^ In recent years, multiple epidemiological studies investigated the association between PPIs use and the risk of preeclampsia.^12,13^ A Swedish study found a reduced risk of preeclampsia among pregnant women who used PPIs in the late maternal stage, while another study from the US found no association of PPIs with decreased risk of preeclampsia.^12,13^ The preliminary search to find any existing meta-analysis was performed on August 2021 in Epistemonikos, PROSPERO, Open Science Framework, Cochrane Library, and JBI Evidence Synthesis. No previous meta-analysis evaluating the risk of preeclampsia among PPIs users were identified. Hence, this study aims to quantify the impact of PPIs use on the risk of preeclampsia in pregnant women.

### Objectives

This meta-analysis aims to assess the risk of preeclampsia among women receiving PPIs during pregnancy.

## Methodology

The study will follow JBI and Cochrane methodology of development of systematic reviews.^14,15^

### Eligibility criteria

We will include all the epidemiological studies assessing the risk of preeclampsia among women receiving PPIs during pregnancy. The current study will be conducted by following this published protocol and will be reported by adhering to the Preferred reporting items for systematic review and meta-analysis (PRISMA) and Meta-analysis of Observational Studies in Epidemiology (<u>MOOSE</u>) reporting guideline.^16,17^

### Characteristics of participants (P)

We will include studies assessing the risk of preeclampsia among pregnant women receiving PPIs anytime during pregnancy.

### Characteristics of exposures (E)

The exposure of interest for the eligible studies will be the use of any PPI including but not limited to omeprazole, esomeprazole, pantoprazole, rabeprazole, lansoprazole, dexlansoprazole, or ilaprazole.

### Characteristics of comparators (C)

We will include all the studies assessing the effect of PPIs on preeclampsia in pregnancy compared to non-PPIs use, or other active drug uses-H2RA antagonist (cimetidine, ranitidine, famotidine, nizatidine, and others).

### Outcomes of interest (O)

The outcome of interest for the eligible studies will be the risk of preeclampsia in pregnant women receiving PPIs.

### Characteristics of study design (S)

Epidemiological analytical studies-cohort studies, case-control studies, longitudinal studies will be eligible for inclusion in this meta-analysis. Animal studies, literature reviews, case reports, *in-vitro* studies, commentary, letters, editorials, and correspondence, will be excluded.

## Information sources and search procedure

### Electronic source and search strategy

A three-step search strategy will be utilized to find both published and unpublished studies. An initial limited search of MEDLINE (Ovid) and Embase (Ovid) will be undertaken, using keywords and index terms related to PPIs and preeclampsia. An analysis of the text words in the title and abstract, and the index terms used to describe the articles will be identified. A second search implementing all identified keywords and index terms will be conducted across all databases of interest. Thirdly, bibliographies of all selected papers will be hand searched, followed by citation tracking to identify any additional articles.

The databases to be searched include MEDLINE (Ovid), Embase (Ovid), Scopus, Web of Science Core Collection, Emcare (Ovid), and CINAHL (EBSCO). Sources of grey literature will be ProQuest Dissertations & Theses Global, and clinical trials register-ClinicalTrials.gov and WHO International Clinical Trials Registry Platform (ICTRP). The article will be searched irrespective of date or language restriction.

### Study selection

All the retrieved articles will be judged based on the PECOS (Population, Exposure, Comparator, Outcomes, and Study design) criteria by two independent reviewers. The study selection process will be completed in two phases. In the first phase, the decision will be made based on information available in the title and abstract. All qualified articles from the first phase will undergo a second screening based on full-text. Any discrepancy regarding study selection will be resolved by discussion between authors. List of excluded studies after full text screening will be developed with reasons for exclusion.

### Assessment of risk of bias

The quality assessment for each study will be performed using Newcastle-Ottawa Scale (NOS) by two reviewers independently. Refer to Appendix I for the items of the NOS.^18^

### Data extraction

Two reviewers will abstract data independently from all the eligible studies. Refer to Appendix II for the sample of the data extraction template. Information based on PECOS criteria will be extracted-study characteristics (study location, study design, study year), population characteristics (patients age, sample size), information on exposure, and study outcomes. In case of insufficient data, primary study authors will be contacted.

## Statistical analysis

Review Manager (RevMan) version 5.4 will be used to perform the meta-analysis using the generic inverse variance method. Risk ratio and odds ratio will be used interchangeably as PPIs use, and preeclampsia events are uncommon events. Therefore, we will use RR representing all these measures for simplicity.^19-21^ Based on Cochrane chi-square value (p< 0.10) and I2 statistics ≥50% meta-analysis model (random effect or fixed effect) will be chosen.^22^ Sensitivity analysis will be performed by the leave-one-out method.^23^ Similarly, subgroup analysis will also be performed based on different comparators, trimester of pregnancy, preterm and term preeclampsia, and others. If meta-analysis is based on ≥10 studies, then a funnel plot will be plotted, and a meta-regression analysis will be conducted.

## Certainty of Evidence

The quality of evidence will be assessed with the Grading of Recommendations Assessment, Development and Evaluation (GRADE) using the five domains: risk of bias, consistency, directness, precision, and publication bias.^24^ The overall certainty of evidence will be considered high (a lot of confidence that the true effect is similar to the estimated effect), moderate (true effect is probably close to the estimated effect), low (true effect might be markedly different from the estimated effect), or very low (true effect is probably markedly different from the estimated effect) for the outcome.

## Data Availability

All data will be provided in the supplementary files.

## Appendix I

### Risk-of-Bias Form

#### NEWCASTLE - OTTAWA QUALITY ASSESSMENT SCALE COHORT STUDIES

<u>Note</u>: A study can be awarded a maximum of one star for each numbered item within the Selection and Outcome categories. A maximum of two stars can be given for Comparability

##### Selection

1. <u>Representativeness of the exposed cohort</u>
  a. truly representative of the general population □
  b. somewhat representative of the general population □
  c. selected group of users eg nurses, volunteers
  d. no description of the derivation of the cohort
2. <u>Selection of the non exposed cohort</u>
  a. drawn from the same community as the exposed cohort □
  b. drawn from a different source
  c. no description of the derivation of the non exposed cohort
3. <u>Ascertainment of exposure</u>
  a. use of accelerometer □
  b. validated questionnaire □
  c. not validated questionnaire or no validation is mentioned
  d. no description
4. <u>Demonstration that outcome of interest was not present at start of study (no asthma at start of study)</u>
  a. Yes □
  b. no

##### Comparability

1. <u>Comparability of cohorts on the basis of the design or analysis</u>
  a. study controls for gender □
  b. study controls for any smoking (eg. parental smoking, past smoking, smoking during pregnancy) AND weight (eg. BMI, overweight, obesity) □

##### Outcome

a. <u>Assessment of outcome</u>
  a. doctor’s diagnosis (not self-reported doctor’s diagnosis) OR objective measurements (eg. Lung function) □
  b. parent/self-reported doctor’s diagnosis OR use of asthma medication □
  c. parent/self-report
  d. no description
b. <u>Was follow-up long enough for outcomes to occur</u>
  a. yes (majority of population at least 1 year) □
  b. no
c. <u>Adequacy of follow up of cohorts</u>
  a. complete follow up - all subjects accounted for □
  b. subjects lost to follow up unlikely to introduce bias - small number lost - > 80 % follow up, or description provided of those lost, proving a non-selective loss to follow up □
  c. follow up rate < 80% and no description of those lost, or a selective loss to follow up
  d. no statement

#### NEWCASTLE - OTTAWA QUALITY ASSESSMENT SCALE CASE CONTROL STUDIES

<u>Note</u>: A study can be awarded a maximum of one star for each numbered item within the Selection and Exposure categories. A maximum of two stars can be given for Comparability.

##### Selection

1. <u>Is the case definition adequate</u>?
  a. yes, with independent validation
  b. yes, eg record linkage or based on self reports
  c. no description
2. <u>Representativeness of the cases</u>
  a. consecutive or obviously representative series of cases
  b. potential for selection biases or not stated
3. <u>Selection of Controls</u>
  a. community controls
  b. hospital controls
  c. no description
4. <u>Definition of Controls</u>
  a. no history of disease (endpoint)
  b. no description of source

##### Comparability

1. <u>Comparability of cases and controls on the basis of the design or analysis</u>
  a. study controls for —————— (Select the most important factor.)
  b. study controls for any additional factor (This criteria could be modified to indicate specific control for a second important factor.)

##### Exposure

1. <u>Ascertainment of exposure</u>
  a. secure record (eg surgical records)
  b. structured interview where blind to case/control status
  c. interview not blinded to case/control status
  d. written self report or medical record only
  e. no description
2. <u>Same method of ascertainment for cases and controls</u>
  a. yes **-**
  b. no
3. <u>Non-Response rate</u>
  a. same rate for both groups
  b. non respondents described
  c. rate different and no designation

## Appendix II

### Data Extraction Form

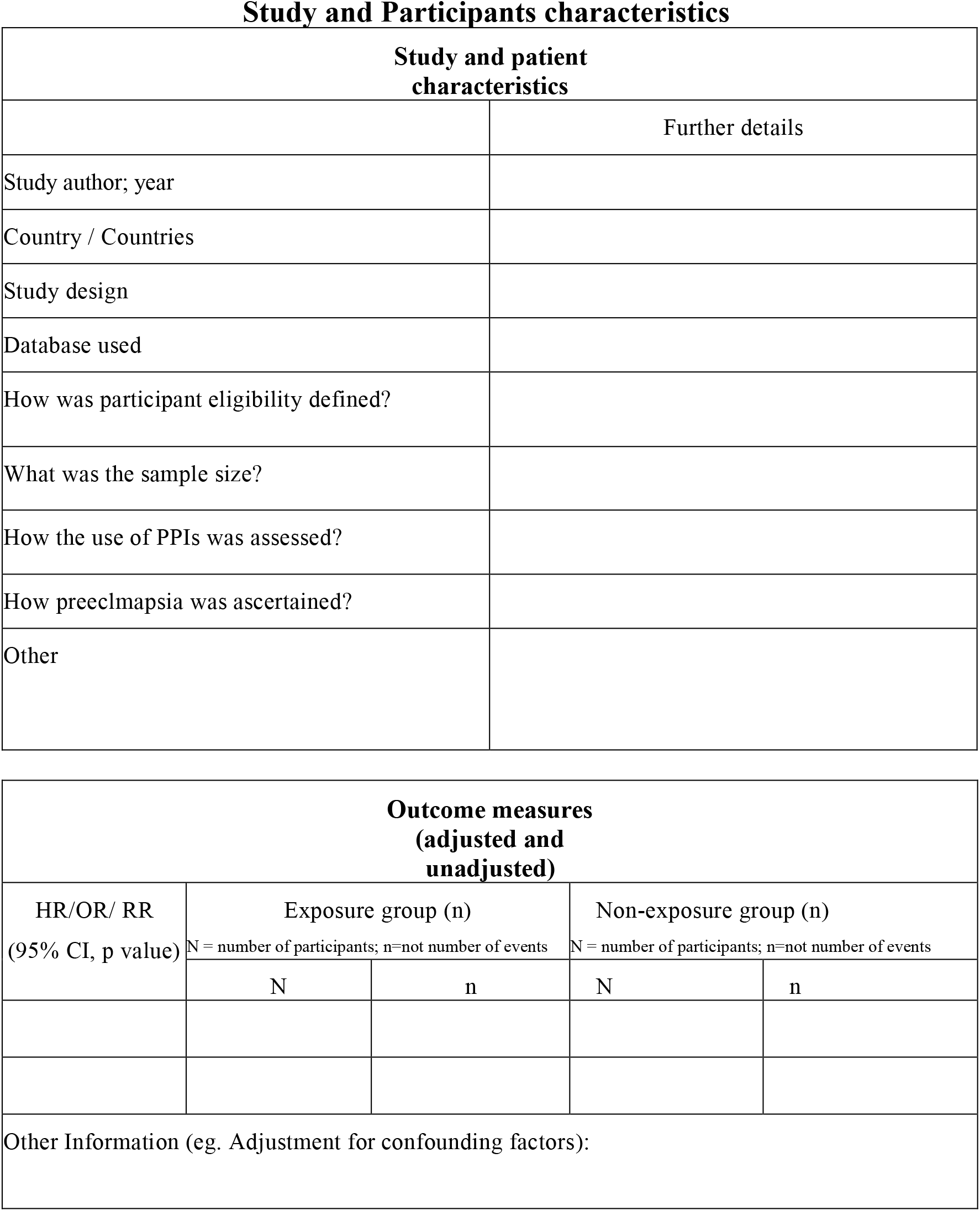

